# Comparative Effectiveness of Dexamethasone in Treatment of Hospitalized COVID-19 Patients during the First Year of the Pandemic: The N3C Data Repository

**DOI:** 10.1101/2022.10.22.22281373

**Authors:** Richard Zhou, Kaitlyn E. Johnson, Justin F. Rousseau, Paul J. Rathouz, The N3C Consortium

## Abstract

**Background:** Dexamethasone, a widely available glucocorticoid, was approved for use in hospitalized COVID-19 patients early in the pandemic based on the RECOVERY trial; however, evidence is still needed to support its real-world effectiveness in patients with a wide range of comorbidities and in diverse care settings.

**Objectives:** To conduct a comparative effectiveness analysis of dexamethasone use with and without remdesivir in hospitalized COVID-19 patients using electronic health record data.

**Methods:** We conducted a retrospective real-world effectiveness analysis using the harmonized, highly granular electronic health record data of the National COVID Cohort Collaborative (N3C) Data Enclave. Analysis was restricted to COVID-19 patients in an inpatient setting, prior to vaccine availability. Primary outcome was in-hospital death; secondary outcome was combined in-hospital death and severe outcome as defined by use of ECMO or mechanical ventilation during stay. Missing data were imputed with single imputation. Matching of dexamethasone-treated patients to non-dexamethasone-treated controls was accomplished using propensity score (PS) matching, stratified by remdesivir treatment and based on demographics, baseline laboratory values, and comorbidities. Treatment benefit was quantified using logistic regression. Further sensitivity analyses were performed using clinical adjusters in matched groups and in strata defined by quartiles of PS.

**Results:** Regression analysis revealed a statistically significant association between dexamethasone use and reduced risk of in-hospital mortality for those not receiving remdesivir (OR=0.77, 95% CI: 0.62 to 0.95, p=0.017), and a borderline statistically significant risk for those receiving remdesivir (OR=0.74, 95% CI: 0.53 to 1.02, p=0.054). Treatment also showed secondary outcome benefit. In sensitivity analyses, treatment effect size generally remained similar with some heterogeneity of benefit across strata of PS.

**Conclusions:** We add evidence that dexamethasone provides benefit with respect to mortality and severe outcomes in a diverse, national hospitalized sample, prior to vaccine availability.

## 1. BACKGROUND AND SIGNIFICANCE

Our aim is to assess the real-world effectiveness of dexamethasone treatment in prevention of poor clinical outcomes among hospitalized COVID-19 patients in the United States (US) during the first year of the pandemic.

At the end of 2020, before vaccinations started to become available, SARS-CoV-2 infection resulted in about 17,000,000 COVID-19 hospitalizations and over 350,000 COVID-19 deaths in the United States alone^1^. In terms of severity, between March and November, 2020, COVID-19 infections resulted in an estimated 12% hospitalization rate, with between 11% and 19% of those experiencing mechanical ventilation, and 9% to 16% mortality^2^. Given the novel emergence of the pandemic and its morbidity and mortality, extraordinary efforts have been undertaken to identify effective treatments for COVID-19. Of great interest since the outset of the pandemic are existing treatments already widely available in hospitals. Especially for severe cases, one such treatment is dexamethasone, a commonly-used glucocorticoid^3^; by one estimate based on 137,870 hospitalized adult COVID-19 patients, 39.1% received dexamethasone during their hospitalization^4^.

Patients with severe COVID-19 exhibit a hyper-inflammatory immune response characterized by elevated proinflammatory cytokines and chemokines^5,6^. Alongside other markers of elevated immune system activity, the hyper-inflammatory immune response contributes to tissue damage and multi-organ failure. A systematic review that considered commonly collected laboratory biomarkers among hospitalized patients identified multiple markers of inflammation and immune system activity, or acute phase reactants, as markedly different among COVID-19 positive (vs negative) hospitalized individuals^7^. Thus, immune system modulation is a critical component of clinical management in hospitalized COVID-19 patients and commonly accomplished through the use of corticosteroids, such as dexamethasone; however, both benefits and poor outcomes have been reported as a result of corticosteroid use^3^.

In June of 2020, the RECOVERY trial, an open-label clinical trial in the United Kingdom, found oral or intravenous (IV) dexamethasone at a dose of 6 mg daily (versus placebo) to be efficacious at reducing the risk of 28-day mortality and duration of hospital stay in patients hospitalized for COVID-19^8^, with an age-adjusted mortality rate ratio of 0.83 (95% CI: 0.75 to 0.93). The lack of equipoise engendered by the RECOVERY trial led to early stopping of three other comparative trials, two of which showed a likely therapeutic benefit for higher dose and intravenous (vs oral) dexamethasone^9,10^, and one of which did not find a benefit for low-dose dexamethasone^11^. After RECOVERY, a meta-analysis of 7 trials in 12 countries and 1703 total critically ill participants showed a favorable odds ratio of 0.66 (95% CI: 0.35 to 0.82) for 28-day mortality comparing systemic administration of corticosteroids to usual care or placebo.

The observational study evidence is recapitulated in a recent meta analysis of 21,350 patients across 73 studies comparing receipt of corticosteroids to no usage^3^. In 8 studies restricted to severe patients, as indicated by mechanical ventilation and/or ICU use, corticosteroids showed a mortality benefit (OR=0.65; 95% CI: 0.51 to 0.83). This is similar to the result of the meta analysis of trial data. However, an additional 32 studies affording comparison had high patient and between-study heterogeneity. These studies suggested a detrimental effect of corticosteroids on mortality (OR=2.30; 95% CI: 1.45 to 3.63); the authors noted that the population for this analysis was highly heterogeneous and selection bias may play a critical and confounding role in this result^3^.

In light of evidence from randomized intervention trials, mixed evidence from observational studies, and current WHO recommendation for use of corticosteroids for severe COVID-19 patients^12^, there remain many questions about use of dexamethasone as front-line clinical therapy for hospitalized COVID-19 patients^13^. This includes confirmatory evidence of effectiveness in real-world settings with a high degree of heterogeneity. This was true at the end of 2020, and improved evidence about this first phase of the pandemic still has important implications for current treatment of hospitalized COVID-19 patients. Such real-world evidence is only possible on a large scale through use of large population-based data repositories, curated to the point of common measures across sources, and sufficiently large sample sizes to support meaningful comparisons. The National COVID Cohort Collaborative (N3C) Data Enclave^14^ was developed to meet this challenge.

Specifically, in the rapidly evolving situation of the COVID-19 pandemic, high quality evidence could not be produced fast enough to guide management decisions. Over two and a half years after the pandemic began, there still remains uncertainty surrounding the effectiveness of first-line treatments for COVID-19. In the absence of infrastructure to rapidly and rigorously evaluate therapeutic interventions across a large enough cohort in diverse settings, the healthcare community made clinical decisions to treat with various interventions of uncertain effectiveness. Individual centers with resources to conduct clinical trials had to generate evidence to drive development of practice guidelines. The health informatics community responded by developing the N3C database as a resource to support rapid generation of evidence through secondary analysis of electronic health record (EHR) data.

## 2. OBJECTIVES

Our aim is to assess the real-world effectiveness of dexamethasone treatment in prevention of poor clinical outcomes among hospitalized COVID-19 patients in the United States (US) during the first year of the pandemic. To accomplish this aim, we leveraged data from the N3C to estimate the real world effectiveness of treatment with dexamethasone in hospitalized COVID-19 patients. In addition to the methodological rigor realized through use of a centralized, harmonized, and highly granular EHR data repository, we demonstrate the usage of data imputation to handle missing biomarkers and a matched propensity score approach to handle biomarkers of disease severity with N3C data to generate high quality evidence. These methods could be applied to a number of similar questions regarding real-world treatment effectiveness within the N3C and beyond.

## 3. METHODS

### 3.1 Study Population and Data

#### 3.1.1 Cohort of hospitalized COVID-19 patients from N3C

The National COVID Cohort Collaborative is a national, representative, and large repository of electronic health record data on COVID-19 cases and controls initiated and developed by the National Institutes of Health (NIH) National Center for Advancing Translational Sciences (NCATS) ^4,14^. The N3C cohort includes patients seen in diverse clinical settings and geographical regions. For this analysis, we used data release version 22 from February 23, 2021, which comprises data on over 65 contributing (single- and multi-site) health systems, over three million COVID-19 positive patients, and over nine million patients in total. The data are harmonized through the Observational Health Data Sciences and Informatics (OHDSI) Observational Medical Outcomes Partnership (OMOP) Common Data Model (CDM). The N3C Data Enclave provides a range of tools to manipulate, transform, analyze and display the data using either written code or graphically-driven commands.

Data for analysis in this report were extracted, manipulated and analyzed in the N3C Data Enclave using Spark SQL (Apache Software Foundation), Python version 3.6 (Python Software Foundation), including PySpark (Apache Software Foundation), and R version 3.5.1 (R Foundation). The N3C’s de-identified dataset was used, which obscures ZIP codes and algorithmically shifts dates of service (while maintaining relative dates of service within each unique cohort member’s trajectory). This project leverages the tables produced in the N3C cohort characterization project ^15^, from which our retrospective cohort and patient variables and severity level were extracted. The overall project is conducted under a Data Use Agreement (DUA) between N3C and the University of Texas (UT) at Austin (PI: PJR) and an N3C-approved Data Use Request (DUR) for this specific study.

#### 3.1.2 Patient and site inclusion criteria and endpoint definition

Our analysis cohort was restricted to adult (over 18 years of age) COVID-19 positive patients with inpatient hospitalizations greater than two days. This specifically excludes emergency department visits which did not result in a hospital admission.

Following Bennett et al. (2021), for primary clinical outcome (endpoint) in the present investigation, patients were classified by their maximum COVID-19 severity level during their hospitalization. We restricted the cohort to three severity levels: *moderate disease*, consisting of inpatient hospitalizations with no use of extracorporeal membrane oxygenation procedure (ECMO) or invasive mechanical ventilation; *severe disease*, consisting of those receiving either ECMO and/or mechanical ventilation; and disease resulting in either in-hospital *death or discharge to hospice*.

The availability of patient laboratory and vital measurements was assessed by provider site. Two sites were removed because there were no laboratory measures available in the data at these sites.

#### 3.1.3 Defining dexamethasone-treated patients and comparison group

From the hospitalized patients matching the patient and site inclusion criteria defined above, we identified the subset of patients who had a record of dexamethasone usage during their selected visit. In that subset, we differentiated the chronic use of dexamethasone for purposes other than treatment of COVID-19 with the use of dexamethasone for the treatment of COVID-19 by first removing those with any dexamethasone administered prior to hospitalization. The patients determined to have been on long-term dexamethasone therapy not for treatment of COVID-19 were not included in either the control or treatment group. We further restricted the entire analysis cohort to (a) those starting dexamethasone treatment in the first two days of hospitalization (treated group), or (b) those who were not treated with dexamethasone during hospitalization (comparison group). Finally, because our study focused on dexamethasone, we removed patients who had records of receiving other corticosteroids (prednisone, methylprednisolone, hydrocortisone) besides dexamethasone from the treatment and comparison groups.

#### 3.1.4 Key variables used in this study

Patient comorbidities, based on conditions in a common comorbidity index (Charlson Comorbidity Index – CCI) ^16^, demographic variables, and laboratory and vital measurement variables were selected based on their availability and clinical significance for use in the imputation, propensity score (PS) matching, and/or logistic regression models predictive of clinical outcome.

A subset of laboratory and vital sign measurement variables were selected from all measurement variables available in the N3C cohort characterization data tables. These variables were selected based on physician expert (JR) opinion of clinical significance. Additionally, selected variables were required to have measurement values available for more than 50% of both the treated and comparison groups at baseline (i.e., taken within ±2 days of visit start). We used a single measurement for all models; in the case of multiple recorded measurement values for a single laboratory test or vital sign, we selected the measurement closest to the visit start for analysis. Before propensity score (PS) matching, variables were less likely to be missing for the treated group than for the comparison group. However, with PS matching, we attempted to balance severity levels between treated group and matched control group, so we included the number (count) of missing laboratory values as an additional matching variable, so that the matched comparison group would have similar missingness patterns as the treated group. As such, after matching, missingness in the comparison group was comparable to that in the treated group.

The following variables were included as adjusters for remaining imbalances in severity at admission in logistic regression models for estimation of treatment effect (see *Treatment effect estimation*, below): age, an updated version of the CCI referred to as Q-score^17^, aspartate transaminase (AST), creatinine, platelet count, and white blood cell count (WBC). The rationale for these last four are that AST and creatinine are indicative of end organ damage (hepatic and renal damage, respectively), while platelet count and WBC reflect inflammatory or infectious response.

For the PS matching model and procedure, the six variables above were included, in addition to the following 13 variables: sex, race, receipt of remdesivir during hospitalization, number of selected laboratory measurements missing before imputation, relative percentage of neutrophils, alanine transaminase (ALT), relative percentage of lymphocytes, albumin, and six comorbidities selected based on association with poor outcomes in hospitalized COVID-19 patients: congestive heart failure (CHF), diabetes mellitus (DM), peripheral vascular disease (PVD), myocardial infarction (MI), pulmonary disease, and cancer. Once these 19 variables were chosen, imputation was performed to allow for matching to patients based on a full dataset, with missing values imputed.

For the imputation procedure, in addition to all above variables, we included the following 7 variables to aid in the prediction of missing values: smoking status, acute kidney injury in hospital defined by change in creatinine from baseline (AKI), ECMO received, mechanical ventilation received, length of hospital stay, body mass index (BMI), and maximum severity level during hospitalization. With the exception of smoking status and BMI, these variables were all indicators of in-hospital outcomes; as such, it is not appropriate to include them as PS matching variables, even while they are valid as a basis for imputation of missing values.

### 3.2 Statistical Analysis

#### 3.2.1 Imputation of missing data

As described, patients, provider sites, and variables were selected to avoid high levels of missingness; however, the analysis dataset still contained a considerable level of missing data, especially for laboratory values. Variables that were never missing by the design of our study include clinical endpoints, treatment with dexamethasone (and also with remdesivir), and provider site. Age was also not ever missing. For comorbidity variables, we assumed that no indication of an existing comorbidity meant the patient was unaffected. Missingness was handled using a multiple imputation (MI) procedure^18^, although owing to limitations in the N3C platform, we generated and analyzed only one imputed dataset. We performed imputation in R using the mice package ^19^ using the 25 variables listed above (*3*.*1*.*4 Key variables used in this study*).

Continuous laboratory measurement variables were first log-transformed and then Winsorized; i.e., values either greater than the 75th percentile plus three-times the IQR, or less than the 25th percentile minus three-times the IQR were shrunk to those two boundaries. Five iterations of the MI algorithm were used to develop and stabilize the conditional “chained equations’’ models for prediction of each potentially missing variable. All other settings were defaults in mice. Continuous variables were imputed using predictive mean matching and the sole categorical variable (race) was imputed using polytomous regression. Quality of imputation was assessed by comparing observed to imputed distributions within each of the treated and comparison groups.

#### 3.2.2 Propensity score matching

In observational data comparative effectiveness investigations, the risk of bias due to treatment assignment being confounded with disease severity, and ultimately clinical endpoint, is always a concern. As commonly done, we employed a propensity score (PS) matching approach to generate a no-corticosteroid comparison group that is closely balanced, in terms of comorbidities and severity at admission, with the dexamethasone treatment group^20^. Propensity score matching was performed in R using the MatchIt package^21^. Separate matching was performed for the treatment group and controls receiving remdesivir and the treatment group and control not receiving remdesivir. Propensity scores were estimated using logistic regression. Model and control units were matched without replacement to dexamethasone-treated units using nearest neighbor matching on the propensity score at a 3:1 (not treated with dex to treated with dex) ratio within the non-remdesivir group and at a 1:1 ratio within the remdesivir group, owing to a smaller number of patients who received remdesivir without dexamethasone. For the remdesivir group, a caliper of 0.65 standard deviation units achieved sufficient balance. The 19 variables included in the PS are listed above (3.*1*.*4 Key variables in this study*). The log-transform of continuous variables at the imputation stage was reversed before inclusion in the PS model. We confirmed that balance in covariates was attained by computing for each covariate the absolute standardized mean difference between the treatment groups; we also compared this difference to that obtained before matching.

#### 3.2.3 Treatment effect estimation

Separately within the pair of matched groups receiving remdesivir and the pair of matched groups not receiving remdesivir, we formally compared the dexamethasone-treated group to the matched control group in two steps. First, we considered analyses stratified by quartile of PS, because the PS is presumably related to disease severity around time of admission, as perceived by the provider(s). For each group, we fitted two logistic regression models with dexamethasone (treatment group) as the primary predictor of interest. The first model considered the combined endpoint of either *severe disease* (see above, *3*.*1*.*2 Patient and site inclusion criteria and endpoint definition*) or *death or hospice referral*, versus *moderate disease*, as the outcome. The second model considered just *death or hospice* (versus *moderate or severe disease*) as the outcome. After having fitted quartile-specific models, in the second step, we fitted an aggregate data model using data from all four quartiles pooled together. For all of these models, we included six adjustors: age, Q-score, AST, creatinine, platelet count, and WBC. The four laboratory values were all log (base-2) transformed; in this way, the regression coefficients will be interpretable as log-odds-ratios of the outcome associated with a 2-fold increase in the (log-base-2 transformed) predictor. As indicated above (see 3.*1*.*4 Key variables used in this study*), the rationale for including these variables, in addition to the wholesale adjustment provided by the PS matched design, is that they were considered a priori to be most strongly predictive of clinical outcome. In addition, these four laboratory values may also have a J- or U-shaped effect. For example, leukocytosis (elevated WBC count) is associated with severe illness/infection, but it is also the case that leukopenia (reduced WBC count) can also be associated with severe illness. As such, we performed sensitivity analyses by re-fitting models including both linear and quadratic versions of the log (base-2) laboratory values.

## 4. RESULTS

### 4.1 Patient Characteristics

Of 4,937 COVID-19 positive patients treated with dexamethasone, 3,645 (82.9%) were inpatient hospitalizations two or more days long where the patient did not also receive prednisone, methylprednisolone, or hydrocortisone. Of these patients, 2,469 received dexamethasone on the first or second day of the visit. After removal of the two sites that had a data quality issue, 2,457 patients remained and were used in imputation and matched to non-dexamethasone treated controls (Figure 1).

**Figure 1.**
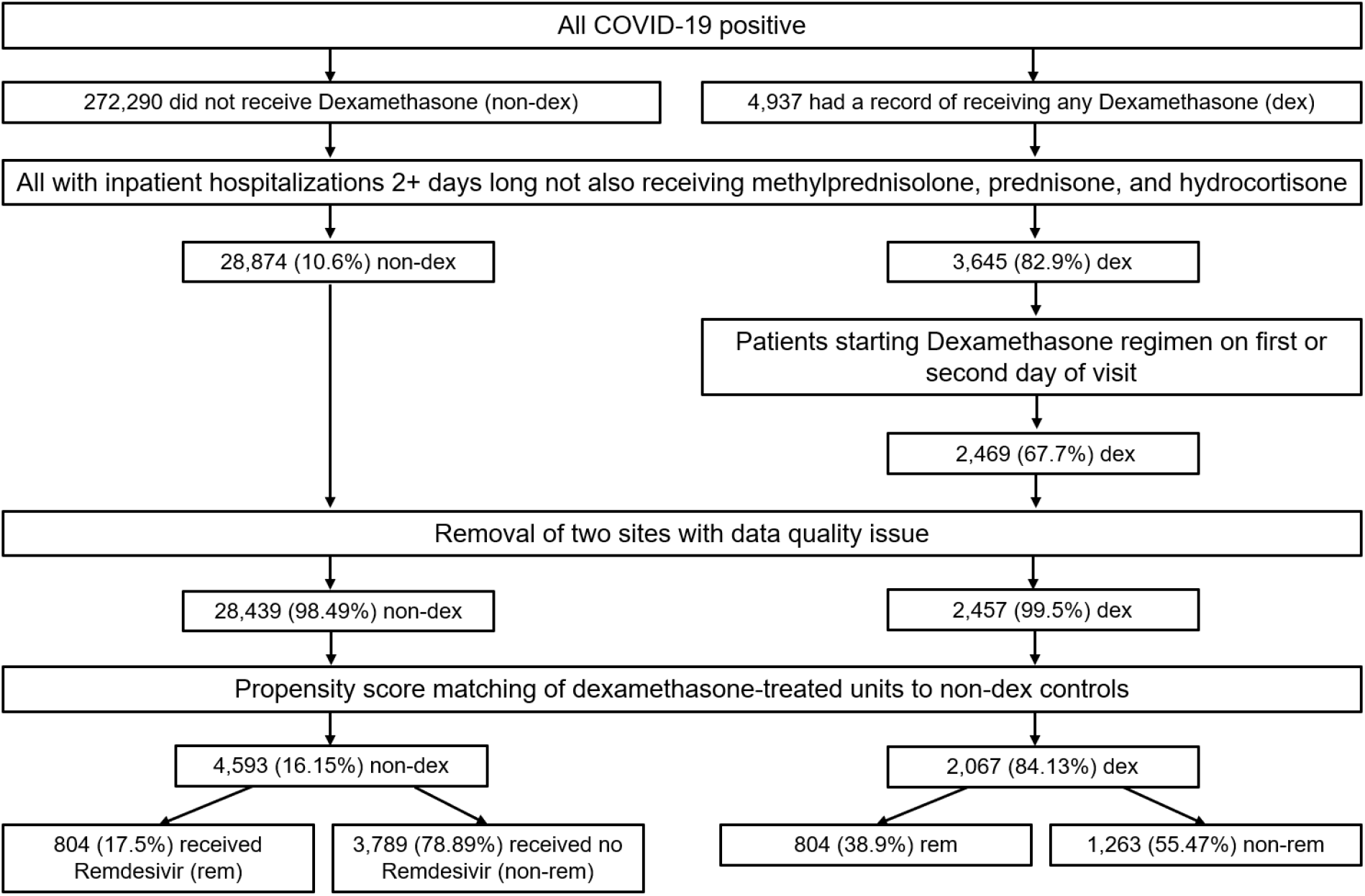
Application of inclusion criteria to patients in the N3C and development of matched pairs for comparing the effect of dexamethasone among patients who had and had not received remdesivir, independently groups. Note that 390 remdesivir and dexamethasone treated patients were dropped between the PS-matching and logistic regression stages because there were not enough controls with propensity scores similar enough to that of the remdesivir and dexamethasone group.

Of 272,290 non-dexamethasone-treated COVID-19 positive patients, 28,874 (10.6%) were inpatient hospitalizations of length two or more days where the patient did not also receive prednisone, methylprednisolone, or hydrocortisone. After removal of the two sites that had a data quality issue, 28,439 patients remained and were used in imputation and as potential controls during PS matching (Figure 1).

The characteristics of the remaining dexamethasone-treated and non-dexamethasone-treated patients after application of the inclusion criteria are shown in Table 1.

**Table 1.**
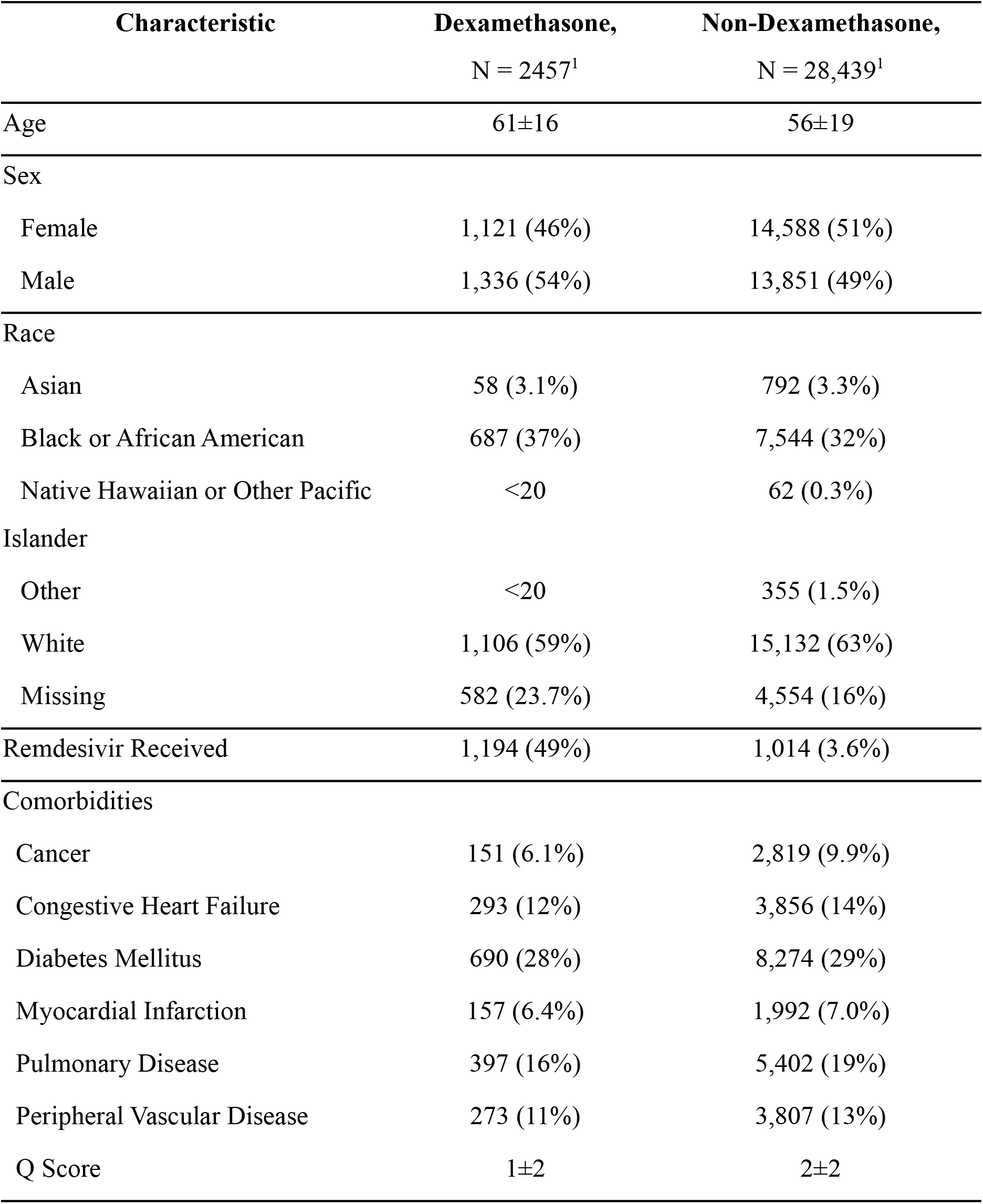
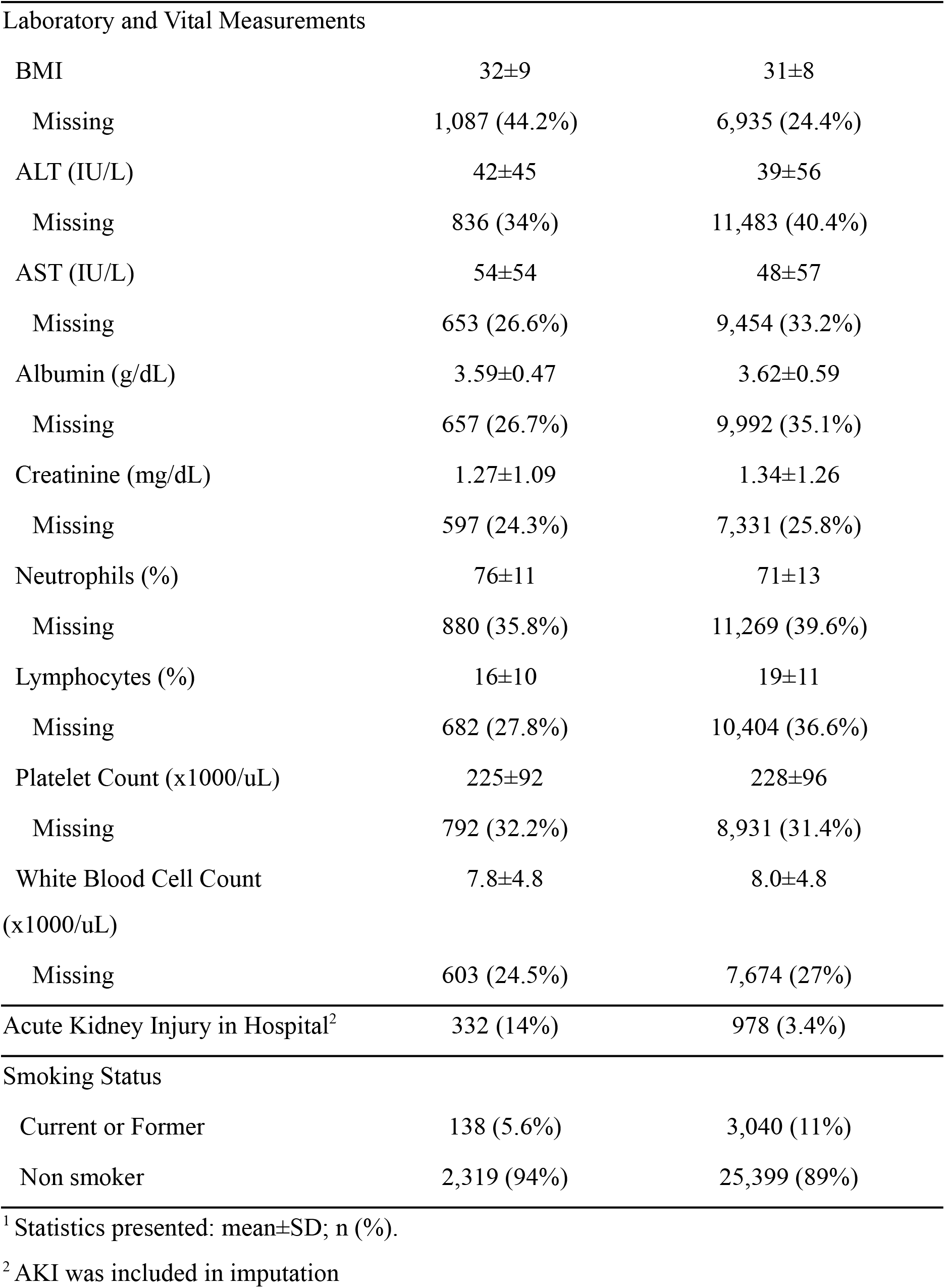
Characteristics of the dexamethasone-treated patients and potential controls at baseline before imputation and PS matching.

### 4.2 Modeling and Outcomes

#### 4.2.1 Imputation and propensity score matching

The percentage of missing values across selected variables is shown in Table 1. The distribution of imputed and observed values for continuous laboratory values was comparable (Appendix A).

Propensity score matching of non-dexamethasone-treated controls to dexamethasone-treated patients in both the remdesivir and non-remdesivir group improved balance. The absolute standardized mean difference between the treated and control group of all continuous covariates and all levels of categorical covariates included in assigning propensity scores was reduced to <0.1 (Appendix B, C). Achieving balance between groups required exclusion of treated units within the dexamethasone and remdesivir treated group which could not be successfully matched to controls. After matching, 2,067 total dexamethasone-treated patients remained. Eight-hundred-four patients were also treated with remdesivir while 1,263 patients were treated with dexamethasone only (Figure 1). Three-hundred-ninety remdesivir and dexamethasone treated patients were dropped between the PS-matching and logistic regression stages because there were not enough controls with propensity scores similar enough to that of the remdesivir and dexamethasone group. In the dexamethasone only group, all units were successfully matched to three controls. The characteristics of the matched groups are summarized in Tables 2a and 2b.

**Table 2a.** Patients not receiving remdesivir: Characteristics of dexamethasone-treated patients and 3:1 PS matched non-dexamethasone-treated controls, after imputation.

| **Characteristic<sup>2</sup>** | **Dexamethasone,**<br>**N = 1263<sup>1</sup>** | **Non-Dexamethasone**<br>**Matched Controls,**<br>**N = 3789<sup>1</sup>** |
| --- | --- | --- |
| Age | 61±16 | 61±17 |
| Sex |  |  |
| Female | 571 (45%) | 1,696 (45%) |
| Male | 692 (55%) | 2,093 (55%) |
| Race |  |  |
| Asian | 52 (4.1%) | 171 (4.5%) |
| Black or African American | 501 (40%) | 1,496 (39%) |
| Native Hawaiian or Other Pacific<br>Islander | <20 | <20 |
| Other | <20 | 48 (1.3%) |
| White | 691 (55%) | 2,059 (54%) |
| Comorbidities |  |  |
| Cancer | 88 (7.0%) | 260 (6.9%) |
| Congestive Heart Failure | 178 (14%) | 523 (14%) |
| Diabetes Mellitus | 379 (30%) | 1,143 (30%) |
| Myocardial Infarction | 104 (8.2%) | 322 (8.5%) |
| Pulmonary Disease | 234 (19%) | 718 (19%) |
| Peripheral Vascular Disease | 157 (12%) | 456 (12%) |
| Q Score | 1±2 | 1±2 |
| Laboratory and Vital Measurements |  |  |
| BMI | 32±9 | 31±8 |
| ALT (IU/L) | 43±57 | 42±58 |
| AST (IU/L) | 56±61 | 56±73 |
| Albumin (g/dL) | 3.57±0.48 | 3.57±0.57 |
| Creatinine (mg/dL) | 1.36±1.23 | 1.36±1.24 |
| Neutrophils (%) | 75±11 | 76±11 |
| Lymphocytes (%) | 16±10 | 16±10 |
| Platelet Count (x1000/uL) | 229±94 | 230±96 |
| White Blood Cell Count<br>(x1000/uL) | 7.9±4.8 | 8.0±4.0 |

<sup>1</sup> Statistics presented: mean±SD; n (%)
<sup>2</sup> Cells with patient count less than 20 obscured to protect confidentiality

**Table 2b.** Patients receiving remdesivir: Characteristics of dexamethasone-treated patients and 1:1 PS matched non-dexamethasone-treated controls, after imputation.

| **Characteristic<sup>2</sup>** | **Dexamethasone,**<br>**N = 804<sup>1</sup>** | **Non-Dexamethasone**<br>**Matched Controls,**<br>**N = 804<sup>1</sup>** |
| --- | --- | --- |
| Age | 62±15 | 62±16 |
| Sex |  |  |
| Female | 365 (45%) | 377 (47%) |
| Male | 439 (55%) | 427 (53%) |
| Race |  |  |
| Asian | <20 | <20 |
| Black or African American | 296 (37%) | 305 (38%) |
| Native Hawaiian or Other Pacific<br>Islander | <20 | <20 |
| Other | <20 | <20 |
| White | 478 (59%) | 464 (58%) |
| Comorbidities |  |  |
| Cancer | 50 (6.2%) | 56 (7.0%) |
| Congestive Heart Failure | 96 (12%) | 102 (13%) |
| Diabetes Mellitus | 242 (30%) | 253 (31%) |
| Myocardial Infarction | 45 (5.6%) | 55 (6.8%) |
| Pulmonary Disease | 138 (17%) | 146 (18%) |
| Peripheral Vascular Disease | 93 (12%) | 94 (12%) |
| Q Score | 1±2 | 1±2 |
| Laboratory and Vital Measurements |  |  |
| BMI | 32±9 | 33±9 |
| ALT (IU/L) | 42±46 | 41±49 |
| AST (IU/L) | 52±52 | 51±44 |
| Albumin (g/dL) | 3.55±0.49 | 3.53±0.52 |
| Creatinine (mg/dL) | 1.18±0.92 | 1.18±0.86 |
| Neutrophils (%) | 75±11 | 75±11 |
| Lymphocytes (%) | 16±10 | 16±9 |
| Platelet Count (x1000/uL) | 221±92 | 222±83 |
| White Blood Cell Count (x1000/uL) | 7.6±4.8 | 7.6±4.5 |

<sup>1</sup> Statistics presented: mean±SD; n (%)
<sup>2</sup> Cells with patient count less than 20 obscured to protect confidentiality

#### 4.2.2 Effect of dexamethasone

Rates of in-hospital death or referral to hospice were lower in the dexamethasone-treated group compared to the non-dexamethasone-treated matched control group for those receiving remdesivir (OR=0.74, 95% CI: 0.53 to 1.02), those not receiving remdesivir (OR=0.77, 95% CI: 0.62 to 0.95), and both remdesivir and non-remdesivir treated groups combined (OR=0.77, 95% CI: 0.64 to 0.91) (Table 3), although the smaller redemdesivir group effect was only borderline significant The use of dexamethasone also was associated with a lower incidence of a combined severe outcome or in-hospital death/hospice referral for those receiving remdesivir (OR=0.82, 95% CI: 0.71 to 0.94), those not receiving remdesivir (OR=0.83, 95% CI: 0.64 to 1.09), and both remdesivir and non-remdesivir treated groups combined (OR=0.84, 95% CI: 0.71 to 1.00) (Table 3).

**Table 3.**
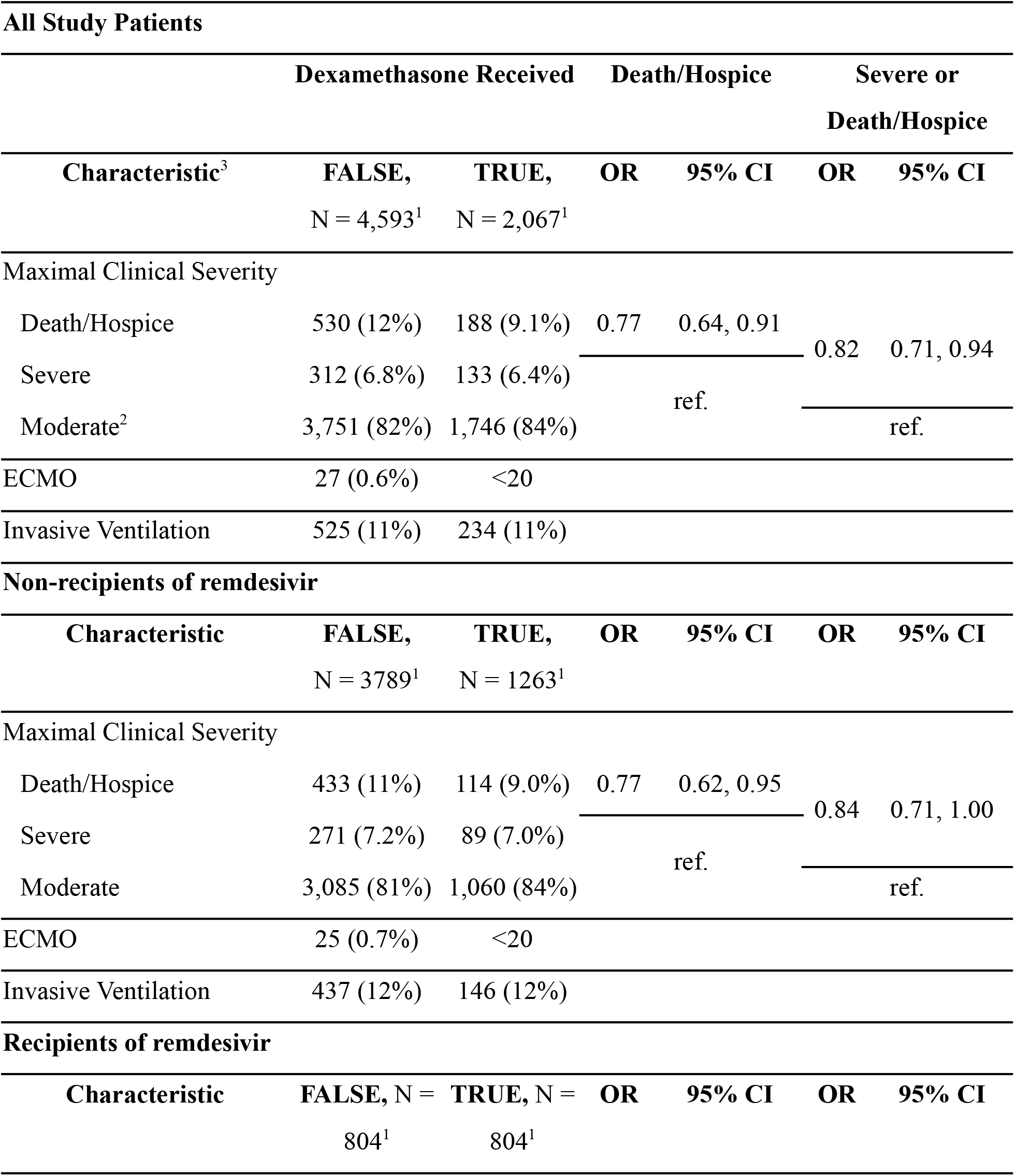
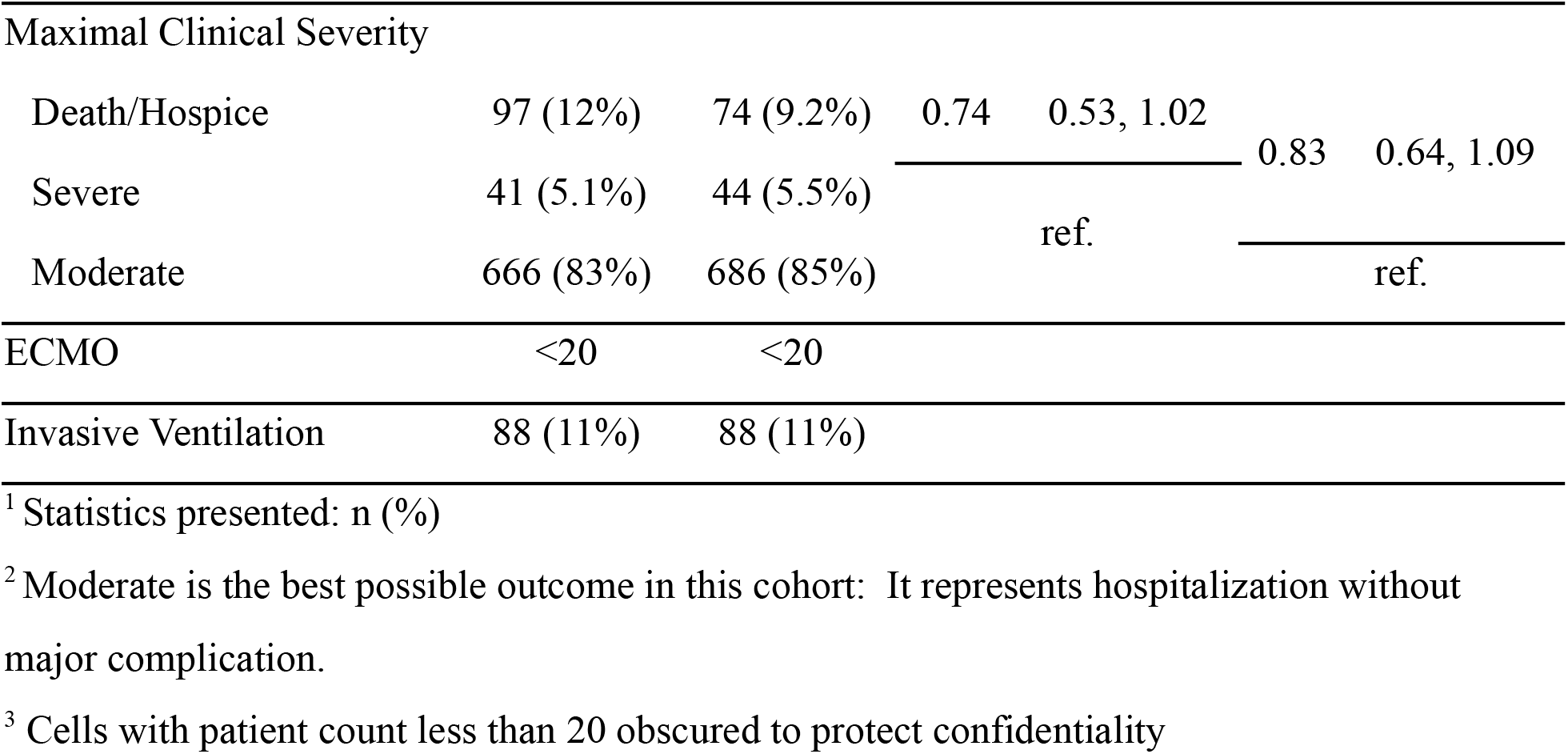
Outcomes and crude odds ratios (95% CI) comparing dexamethasone recipients to PS-matched non-recipients in N3C hospitalized COVID-19 patients. Non-recipients of remdesivir are matched 3:1 (non-dex to dex), while remdesivir recipients are matched 1:1.

In sensitivity analyses, the effect of dexamethasone on reduction of in-hospital death/hospice referral (OR=0.74, 95% CI: 0.59 to 0.93, p=0.012) and combined severe outcome or in-hospital death/hospice referral (OR=0.81, 95% CI: 0.67 to 0.96, p=0.02) in the non-remdesivir group remained similar and significant after adjusting for the effect of age, Q-score, AST, creatinine, platelet count, and white blood cell count (Table 4a). Within the strata defined by quartiles of propensity score, the effect of dexamethasone on reduction of both study outcomes in the non-remdesivir group after adjustment was stronger in the fourth quartile of propensity scores (presumably patients who presented as most severe) than in the aggregate cohort (OR=0.62, 95% CI: 0.42 to 0.9, p=0.014 mortality outcome; OR=0.61, 95% CI: 0.44 to 0.82, p=0.002, combined severe/mortality outcome) (Appendix D, E). In the first through third quartiles, the effect was non-significant and somewhat heterogeneous.

**Table 4a.** Patients not receiving remdesivir: Prediction of in-hospital death/hospice referral and combined in-hospital death/hospice referral and severe outcome by receipt of dexamethasone with logistic regression models.

| **Aggregate PS Matched Cohort<sup>4</sup>** |  |  |  |  |  |  |
| --- | --- | --- | --- | --- | --- | --- |
| **Characteristic<sup>2,3,4</sup>** | **Death/Hospice** |  |  | **Severe or Death/Hospice** |  |  |
|  | **OR<sup>1</sup>** | **95% CI<sup>1</sup>** | **p-value** | **OR<sup>1</sup>** | **95% CI<sup>1</sup>** | **p-value** |
| Dexamethasone | 0.75 | 0.59, 0.95 | 0.017 | 0.82 | 0.68, 0.98 | 0.028 |
| Age | 1.06 | 1.05, 1.07 | <0.001 | 1.03 | 1.02, 1.03 | <0.001 |
| Q-Score | 1.14 | 1.09, 1.18 | <0.001 | 1.11 | 1.07, 1.15 | <0.001 |
| AST | 1.34 | 1.22, 1.47 | <0.001 | 1.37 | 1.27, 1.47 | <0.001 |
| Creatinine | 1.27 | 1.14, 1.42 | <0.001 | 1.21 | 1.10, 1.32 | <0.001 |
| Platelet | 0.64 | 0.54, 0.76 | <0.001 | 0.69 | 0.60, 0.79 | <0.001 |
| WBC | 1.89 | 1.62, 2.21 | <0.001 | 2.01 | 1.77, 2.28 | <0.001 |

<sup>1</sup> OR = Odds Ratio, CI = Confidence Interval.
<sup>2</sup> The model included a categorical variable indicating which PS quartile the patient was in (Q1, Q2, Q3, Q4)
<sup>3</sup> AST, creatinine, platelet count, and WBC count were log-base-2 transformed.
<sup>4</sup> See Appendix D for results within strata defined by quartile of PS

In the remdesivir-treated group, the use of dexamethasone showed a statistically significant benefit in reducing in-hospital death or referral to hospice (OR=0.71; 95% CI: 0.51 to 1.00, p=0.048); however, the effect of dexamethasone on the combined severe or in-hospital death/referral to hospice outcome was non-significant (Table 4b).

**Table 4b.**
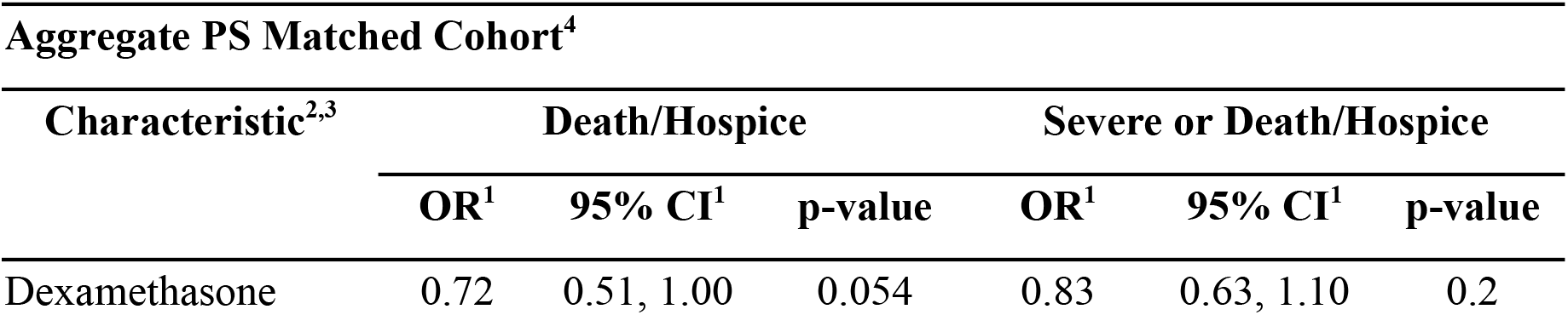
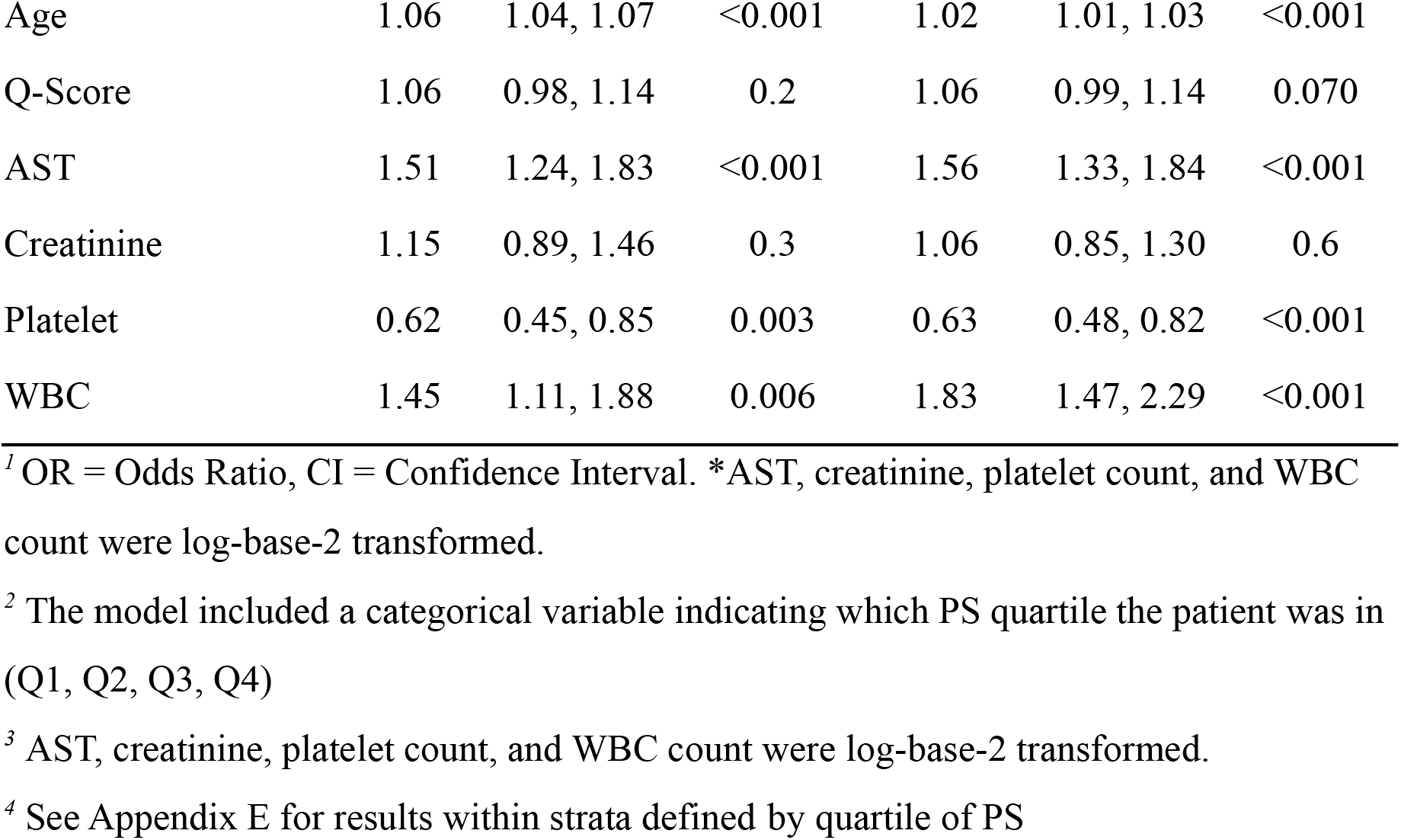
Patients receiving remdesivir: Prediction of in-hospital death/hospice referral and combined in-hospital death/hospice referral and severe outcome by receipt of dexamethasone with logistic regression models.

| **Aggregate PS Matched Cohort<sup>4</sup>** |  |  |  |  |  |  |
| --- | --- | --- | --- | --- | --- | --- |
| **Characteristic<sup>2,3</sup>** | **Death/Hospice** |  |  | **Severe or Death/Hospice** |  |  |
|  | **OR<sup>1</sup>** | **95% CI<sup>1</sup>** | **p-value** | **OR<sup>1</sup>** | **95% CI<sup>1</sup>** | **p-value** |
| Dexamethasone | 0.72 | 0.51, 1.00 | 0.054 | 0.83 | 0.63, 1.10 | 0.2 |
| Age | 1.06 | 1.04, 1.07 | <0.001 | 1.02 | 1.01, 1.03 | <0.001 |
| Q-Score | 1.06 | 0.98, 1.14 | 0.2 | 1.06 | 0.99, 1.14 | 0.070 |
| AST | 1.51 | 1.24, 1.83 | <0.001 | 1.56 | 1.33, 1.84 | <0.001 |
| Creatinine | 1.15 | 0.89, 1.46 | 0.3 | 1.06 | 0.85, 1.30 | 0.6 |
| Platelet | 0.62 | 0.45, 0.85 | 0.003 | 0.63 | 0.48, 0.82 | <0.001 |
| WBC | 1.45 | 1.11, 1.88 | 0.006 | 1.83 | 1.47, 2.29 | <0.001 |

<sup>1</sup> OR = Odds Ratio, CI = Confidence Interval. \*AST, creatinine, platelet count, and WBC count were log-base-2 transformed.
<sup>2</sup> The model included a categorical variable indicating which PS quartile the patient was in (Q1, Q2, Q3, Q4)
<sup>3</sup> AST, creatinine, platelet count, and WBC count were log-base-2 transformed.
<sup>4</sup> See Appendix E for results within strata defined by quartile of PS

The same benefit of dexamethasone for both the remdesivir and non-remdesivir groups was observed in further sensitivity analyses where the logistic regression models above with linear adjusters were extended to investigate quadratic effects for the laboratory variables included, though the effect of dexamethasone on the combined outcome was again not significant in the remdesivir group (Appendix F, G).

## 5. DISCUSSION

Our analysis of multi-site EHR data in the first year of the pandemic confirmed existing clinical trial findings that dexamethasone shows in-hospital mortality benefit. We also found that use of dexamethasone results in a reduction in the secondary outcome of in-hospital mortality or severe outcome defined by the use of ECMO or mechanical ventilation. The effect of dexamethasone on reducing the combined severe and in-hospital death/hospice referral outcome was not as strong as the effect of dexamethasone on in-hospital death/hospice referral alone, although this difference was relatively modest. Similar outcomes were observed in both the remdesivir-treated and non-remdesivir-treated groups.

Whereas evidence for benefit of dexamethasone was shown in the RECOVERY trial, real-world effectiveness of dexamethasone over a wide range of patients with varying levels of comorbidities and baseline severity was needed to ensure that its benefit applied in these settings. Meta-analyses of corticosteroid trials and studies showed general benefit, but mixed results. Additionally, there were many clinical questions concerning the use of dexamethasone and corticosteroids, of which many remain. Although co-administration of remdesivir with dexamethasone was not directly assessed by our study, our analysis suggests that remdesivir did not appear to affect the benefit of dexamethasone in reduction of either mortality or the secondary outcome.

The treatment effects observed in the cohorts were subjected to sensitivity analyses. Treatment effect size was the same when checking for both linear and quadratic effects of included adjusters; however, we found that the odds ratios for the secondary outcome in the remdesivir groups were non-significant. It is likely that more data are needed in the remdesivir groups to confirm the effect. Sensitivity analyses also involved assessment of outcome by quartile of propensity score. In both remdesivir and non-remdesivir groups, there was significant variability of the odds ratios across strata; however, one notable trend was a statistically significant reduction in both study outcomes in the fourth quartile of the non-remdesivir analysis group. This suggests those in the fourth quartile of PS, who were likely perceived as most severe at baseline, benefited most from dexamethasone treatment in comparison to other patients assigned dexamethasone treatment, consistent with current practice of use of dexamethasone primarily in more severe patients.

Several issues related to the statistical methodology arose in the course of our study, and some comments about those follow here. First, we expected to find very few patients who had received remdesivir in the absence of dexamethasone or another corticosteroid. Indeed, this number was not large, but it did allow for comparison of dexamethasone treatment to lack thereof within a separate remdesivir stratum – we did not feel that remdesivir recipients and non-recipients could be combined in primary analyses, although we do present some pooled results for ease of exposition. Nevertheless, it could be that within the remdesivir stratum, the PS-based matching on severity is less complete than in the non-remdesivir stratum. The two strata do in principle allow for comparison of dexamethasone treatment effects between those also receiving and not receiving remdesivir. We did not detect strong evidence of such heterogeneity of treatment effects, although our sample size was limited to be able to detect such interaction terms.

Second, regarding concerns about the considerable amount of missing data, we note that other N3C investigators have also struggled with this issue and have nevertheless come away able to draw robust and useful conclusions using the N3C platform. A major strength of our analysis is that we matched on the number of missing lab values as a predictor of dexamethasone treatment (versus no such treatment). Of course, we counted the number of missing values before imputation, and then conducted imputation before running the PS matching algorithm. Indeed, those treated with dexamethasone had far more complete laboratory profiles, most likely reflecting severity of disease. This component of matching may be one of the more powerful variables to achieve balance on severity (which is not directly observed).

A third issue is that we were only able to perform a single imputation of missing values (i.e., a single iteration of a standard multiple imputation algorithm). This is of little consequence in terms of effect estimation, but it could lead to mild underestimation of standard errors, and this is a limitation. However, the architecture of the N3C platform meant that the imputation, the PS matching, and the logistic regression analyses each took place in their own N3C “data transformation” nodes. In order to conduct multiple imputation, we would need to create multiple parallel pipelines across such nodes, a task beyond our capabilities. Despite these issues, the benefit of the comprehensiveness and national representativeness of the N3C platform outweighed these methodological considerations.

There are also some other general limitations associated with the data to be aware of. These are mostly fundamental limitations that come with the use of electronic health record data, especially data harmonized across many sites. Underlying causes of missing data are not always clear. Likewise, the sites contributing to the N3C may not be an accurate representation of the population, as there may be selection bias in which sites have the resources to be able to contribute to the N3C database. Additionally, there is uncertainty in available measurements and the details of the measurement procedure at each source – and how such procedures vary across sources – are generally not available. These issues may result in bias, though we did not notice any abnormal missingness patterns and we detected and handled implausible measurement values in the statistical analysis. Patients were assumed to be unaffected if a given comorbidity was not present in the data, though it was reported that around half of patients in the N3C have pre-existing health condition data available to allow determination of comorbidity^15^. Demographic data such as race have known quality limitations when collected from EHRs which may contribute to bias^22^. Key data such as detailed ventilator flow settings, ICU admission, oxygen saturation (SpO_2_), and supplemental oxygen that would have enabled us to answer additional questions about the effects of dexamethasone were also not available in the N3C. Finally, specific details about the administration of dexamethasone including delivery route (IV vs oral) and dosage were absent in a portion of patients who received dexamethasone, limiting the potential to study how these factors specifically affect outcomes.

Despite limitations which make the exploration of certain questions less feasible, the large number of sites and patient-level variables included in the N3C enabled our study of dexamethasone on patient outcome and interaction with remdesivir, adding to the body of evidence that dexamethasone generally reduces mortality and severe outcomes whether administered alone or in addition to remdesivir. There is great potential to use the N3C for future research in related directions. The effect of other corticosteroids or drugs which were used or considered to treat hospitalized COVID-19 can be studied in a similar way as described in this work. More drug-drug interactions may be explored as the data resource grows. Additionally, future research could include subgroup analyses to answer questions about the effect of certain drugs on outcomes within more specific groups of interest or even uncover groups with different outcomes defined by, for example, key biomarkers.

As the N3C resource continues to improve the quality, quantity, and veracity of data, increase the diversity of sites contributing, and expand the granularity of data elements, the potential for higher quality analyses increases. We demonstrated the value of the N3C as a resource with the use of methods described in this paper to conduct robust secondary analyses of EHR data producing high quality evidence evaluating the effectiveness of interventions to manage COVID-19. This provides a framework to equip the field to respond quickly to generate evidence to guide management interventions when facing the next emerging, rapidly evolving pandemic when there is not sufficient time to conduct robust prospective clinical trials.

## 6. CONCLUSIONS

Using real-world data for effectiveness analysis of dexamethasone therapy in hospitalized COVID-19 patients prior to vaccine availability, this study on a national EHR sample adds evidence that such therapy provides benefit with respect to mortality and severe outcomes such as mechanical ventilation or ECMO.

### Clinical Relevance Statement

Our real-world effectiveness analysis supports dexamethasone as an option for hospitalized COVID-19 patients, useful in reduction of mortality and severe outcomes, in a heterogeneous group of patients and matched, untreated, controls. We find weaker but positive support for dexamethasone in combination with remdesivir versus remdesivir alone. We also find evidence that the most severe patients at baseline may benefit most from dexamethasone treatment.

## Data Availability

The N3C Data Enclave (covid.cd2h.org/enclave) houses the fully reproducible, transparent, and broadly available limited and de-identified datasets (HIPAA definitions: https://www.hhs.gov/hipaa/for-professionals/privacy/special-topics/de-identification/index.html) used in this project. Data is accessible by investigators at institutions that have signed a Data Use Agreement with NIH who have taken human subjects and security training and attest to the N3C User Code of Conduct. Investigators wishing to access the limited dataset must also supply an institutional IRB protocol. All requests for data access are reviewed by the NIH Data Access Committee. A full description of the N3C Enclave governance has been published; information about how to apply for access is available on the NCATS website: https://ncats.nih.gov/n3c/about/applying-for-access. Reviewers and health authorities will be given access permission and guidance to aid reproducibility and outcomes assessment. A Frequently Asked Questions about the data and access has been created at https://ncats.nih.gov/n3c/about/program-faq.
The code for this study (DUR: RP-11698A) is located within the N3C Data Enclave and can be accessed upon request, provided the user already has access to the N3C.

https://zenodo.org/communities/cd2h-covid/

https://github.com/National-COVID-Cohort-Collaborative

## Acknowledgements

This study was funded by NIH Grant R01 AI151176 (KJ), CDC Grant U01IP001136 (KJ), core funds of the Dell Medical School of the University of Texas at Austin (PJR), and in part by a donation from Tito’s Handmade Vodka (RZ, PJR).

The analyses described in this publication were conducted with data or tools accessed through the NCATS N3C Data Enclave (https://covid.cd2h.org) and N3C Attribution & Publication Policy v1.2-2020-08-25b supported by NCATS U24 TR002306. This research was possible because of the patients whose information is included within the data and the organizations (https://ncats.nih.gov/n3c/resources/data-contribution/data-transfer-agreement-signatories) and scientists who have contributed to the on-going development of this community resource ^14^.

## Disclaimer

The content is solely the responsibility of the authors and does not necessarily represent the official views of the National Institutes of Health or the N3C program.

We gratefully acknowledge the following core contributors to N3C: Adam B. Wilcox, Adam M. Lee, Alexis Graves, Alfred (Jerrod) Anzalone, Amin Manna, Amit Saha, Amy Olex, Andrea Zhou, Andrew E. Williams, Andrew Southerland, Andrew T. Girvin, Anita Walden, Anjali A. Sharathkumar, Benjamin Amor, Benjamin Bates, Brian Hendricks, Brijesh Patel, Caleb Alexander, Carolyn Bramante, Cavin Ward-Caviness, Charisse Madlock-Brown, Christine Suver, Christopher Chute, Christopher Dillon, Chunlei Wu, Clare Schmitt, Cliff Takemoto, Dan Housman, Davera Gabriel, David A. Eichmann, Diego Mazzotti, Don Brown, Eilis Boudreau, Elaine Hill, Elizabeth Zampino, Emily Carlson Marti, Emily R. Pfaff, Evan French, Farrukh M Koraishy, Federico Mariona, Fred Prior, George Sokos, Greg Martin, Harold Lehmann, Heidi Spratt, Hemalkumar Mehta, Hongfang Liu, Hythem Sidky, J.W. Awori Hayanga, Jami Pincavitch, Jaylyn Clark, Jeremy Richard Harper, Jessica Islam, Jin Ge, Joel Gagnier, Joel H. Saltz, Joel Saltz, Johanna Loomba, John Buse, Jomol Mathew, Joni L. Rutter, Julie A. McMurry, Justin Guinney, Justin Starren, Karen Crowley, Katie Rebecca Bradwell, Kellie M. Walters, Ken Wilkins, Kenneth R. Gersing, Kenrick Dwain Cato, Kimberly Murray, Kristin Kostka, Lavance Northington, Lee Allan Pyles, Leonie Misquitta, Lesley Cottrell, Lili Portilla, Mariam Deacy, Mark M. Bissell, Marshall Clark, Mary Emmett, Mary Morrison Saltz, Matvey B. Palchuk, Melissa A. Haendel, Meredith Adams, Meredith Temple-O’Connor, Michael G. Kurilla, Michele Morris, Nabeel Qureshi, Nasia Safdar, Nicole Garbarini, Noha Sharafeldin, Ofer Sadan, Patricia A. Francis, Penny Wung Burgoon, Peter Robinson, Philip R.O. Payne, Rafael Fuentes, Randeep Jawa, Rebecca Erwin-Cohen, Rena Patel, Richard A. Moffitt, Richard L. Zhu, Rishi Kamaleswaran, Robert Hurley, Robert T. Miller, Saiju Pyarajan, Sam G. Michael, Samuel Bozzette, Sandeep Mallipattu, Satyanarayana Vedula, Scott Chapman, Shawn T. O’Neil, Soko Setoguchi, Stephanie S. Hong, Steve Johnson, Tellen D. Bennett, Tiffany Callahan, Umit Topaloglu, Usman Sheikh, Valery Gordon, Vignesh Subbian, Warren A. Kibbe, Wenndy Hernandez, Will Beasley, Will Cooper, William Hillegass, Xiaohan Tanner Zhang. Details of contributions available at covid.cd2h.org/core-contributors.

## Conflict of Interest

The authors declare they have no conflict of interest in this research.

## Human Subjects Protections

The study was determined to be “not human subjects research” by the University of Texas Institutional Review Board.

## Data/Code Availability Statement

The N3C Data Enclave (covid.cd2h.org/enclave) houses the fully reproducible, transparent, and broadly available limited and de-identified datasets (HIPAA definitions: https://www.hhs.gov/hipaa/for-professionals/privacy/special-topics/de-identification/index.html) used in this project. Data is accessible by investigators at institutions that have signed a Data Use Agreement with NIH who have taken human subjects and security training and attest to the N3C User Code of Conduct. Investigators wishing to access the limited dataset must also supply an institutional IRB protocol. All requests for data access are reviewed by the NIH Data Access Committee. A full description of the N3C Enclave governance has been published; information about how to apply for access is available on the NCATS website: https://ncats.nih.gov/n3c/about/applying-for-access. Reviewers and health authorities will be given access permission and guidance to aid reproducibility and outcomes assessment. A Frequently Asked Questions about the data and access has been created at https://ncats.nih.gov/n3c/about/program-faq. Additionally, please see the below links for further information:

https://github.com/National-COVID-Cohort-Collaborative

https://zenodo.org/communities/cd2h-covid/

The code for this study (DUR: RP-11698A) is located within the N3C Data Enclave and can be accessed upon request, provided the user already has access to the N3C.

## Notes

### Competing Interest Statement

The authors have declared no competing interest.

### Author Declarations

The University of Texas Institutional Review Board waived ethical approval for this work; the study was determined to be "not human subjects research".

