## Appendices for "Comparative Effectiveness of Dexamethasone in Treatment of Hospitalized COVID-19 Patients during the First Year of the Pandemic: The N3C Data Repository"

**Appendix A.** Imputed versus observed distributions for imputed continuous variables.

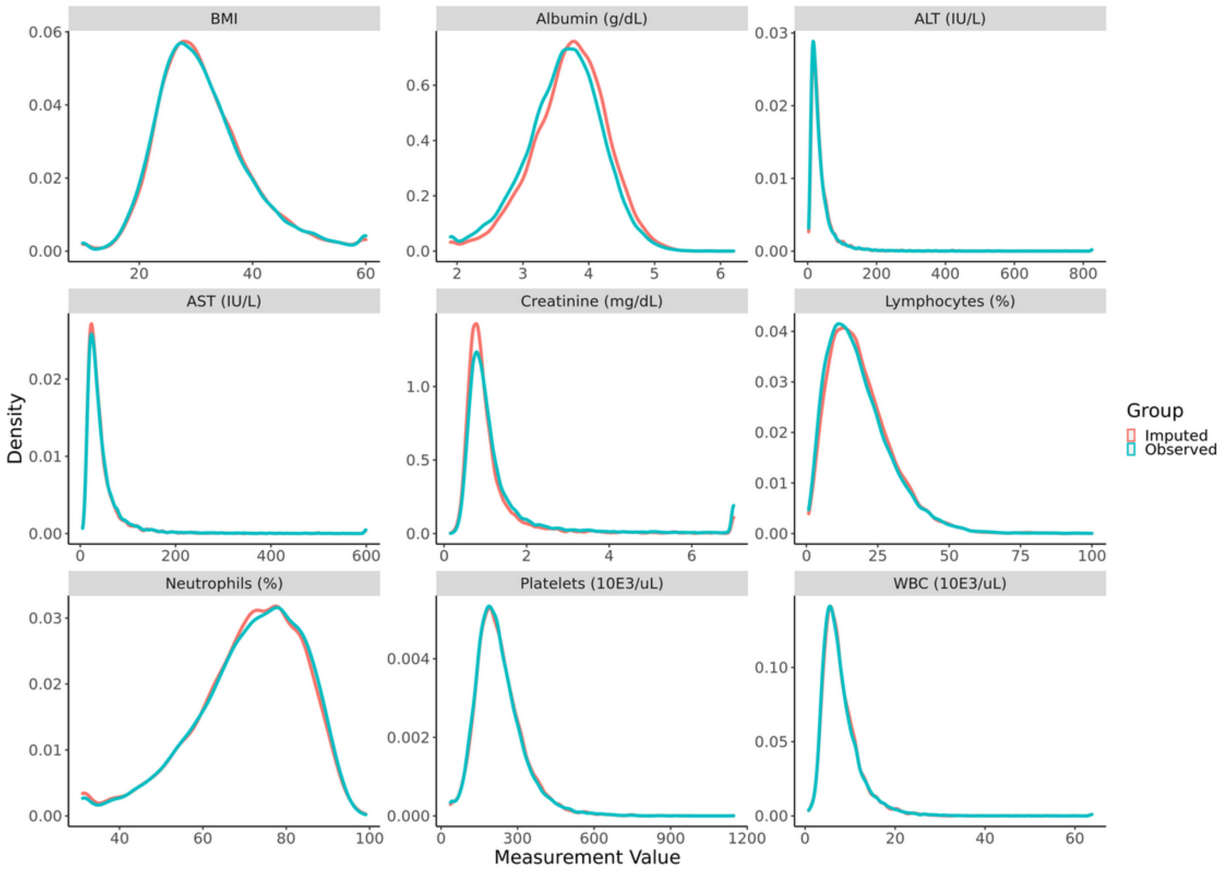

**Appendix B.** Absolute standardized mean difference before and after 3:1 propensity score matching of non-dexamethasone controls to dexamethasone treated patients. All patients did not receive remdesivir.

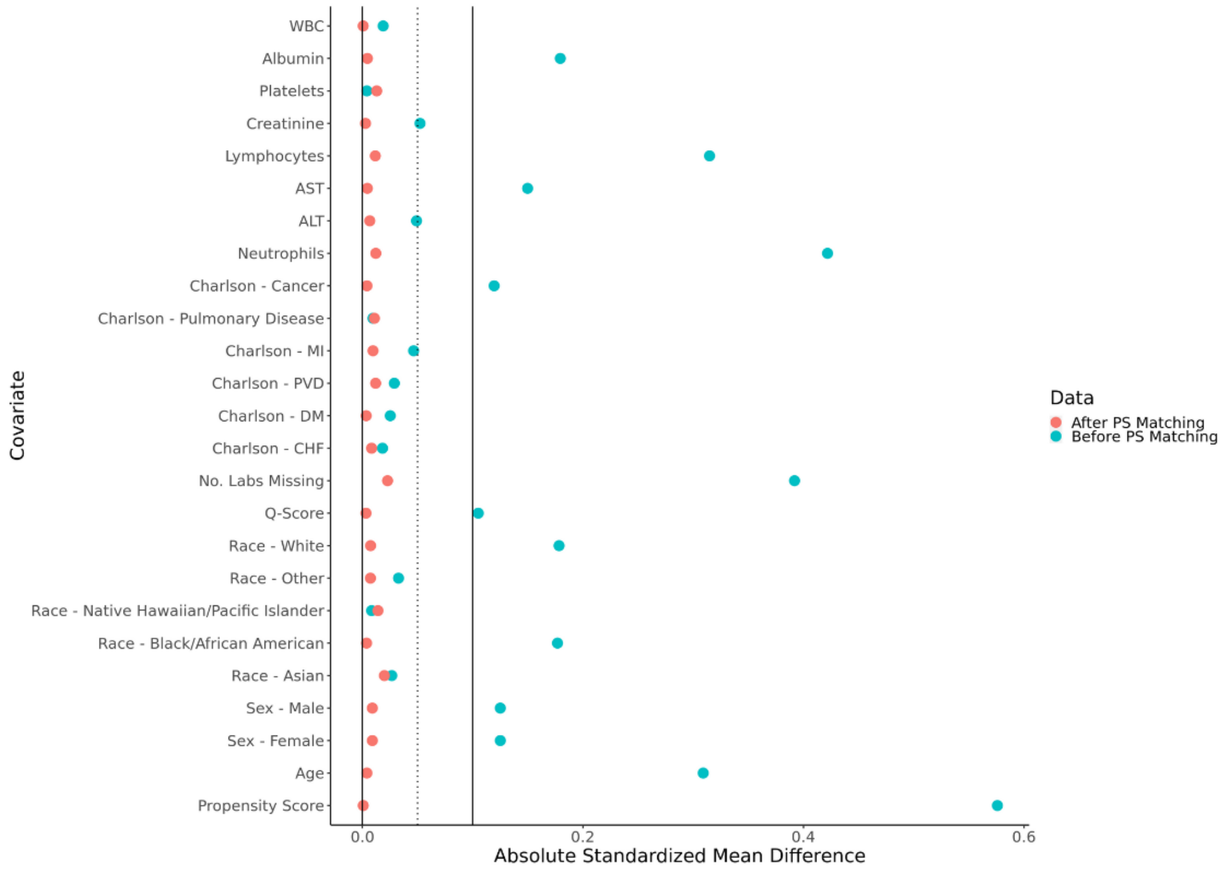

**Appendix C.** Absolute standardized mean difference before and after 1:1 propensity score matching of non-dexamethasone controls to dexamethasone treated patients. All patients received remdesivir.

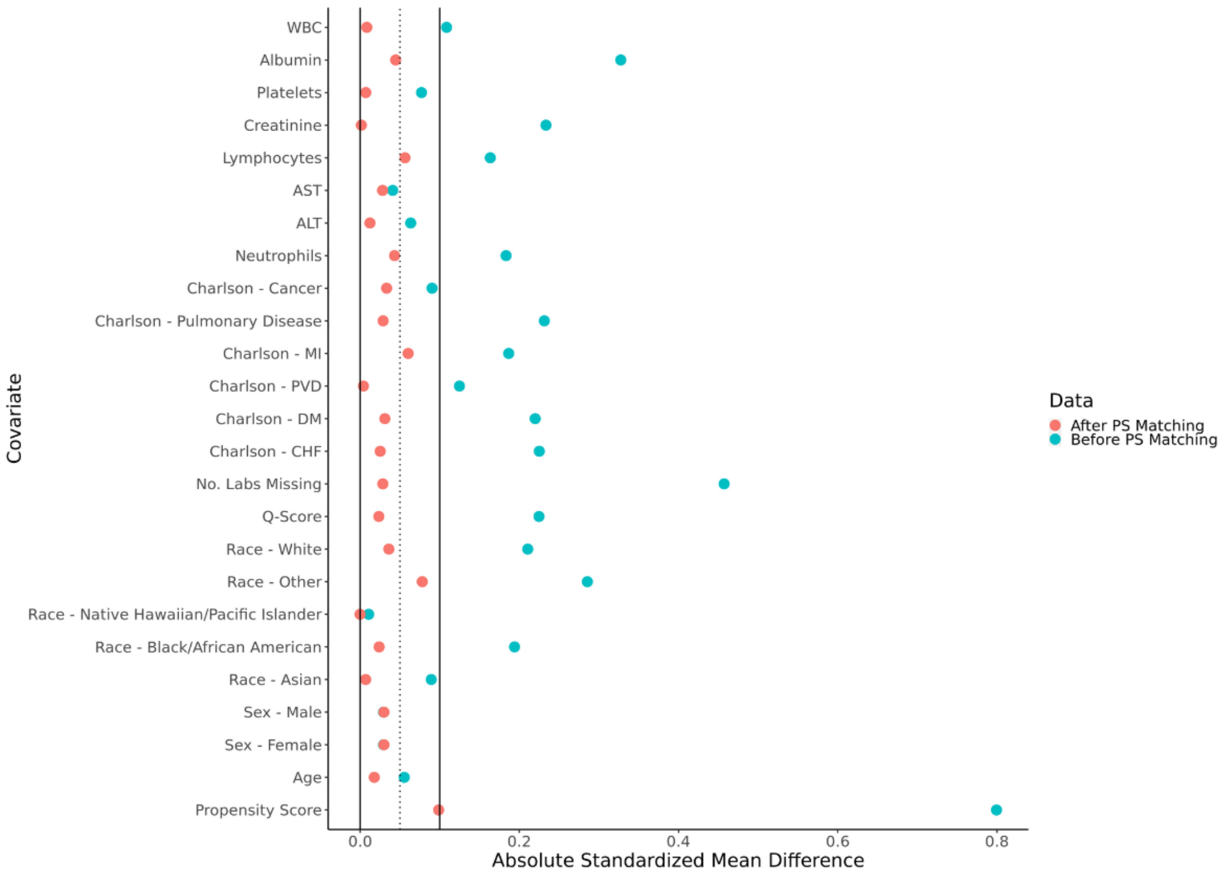

**Appendix D.** Patients not receiving remdesivir: Prediction of in-hospital death/hospice referral and combined in-hospital death/hospice referral and severe outcome by receipt of dexamethasone with logistic regression models, by strata of PS.

| Characteristic* | Death/Hospice | | | Severe or Death/Hospice | | |
|---|---|---|---|---|---|---|
| | OR[1] | 95% CI[1] | p-value | OR[1] | 95% CI[1] | p-value |
| **1st Quartile PS** | | | | | | |
| Dexamethasone | 0.59 | 0.32, 1.06 | 0.089 | 0.86 | 0.54, 1.33 | 0.5 |
| Age | 1.06 | 1.04, 1.08 | <0.001 | 1.04 | 1.03, 1.05 | <0.001 |
| Q-Score | 1.14 | 1.06, 1.22 | <0.001 | 1.13 | 1.06, 1.20 | <0.001 |
| AST | 0.83 | 0.61, 1.11 | 0.2 | 1.16 | 0.94, 1.44 | 0.2 |
| Creatinine | 1.25 | 0.96, 1.60 | 0.093 | 1.09 | 0.87, 1.35 | 0.4 |
| Platelets | 0.47 | 0.32, 0.69 | <0.001 | 0.60 | 0.44, 0.83 | 0.002 |
| WBC | 2.16 | 1.57, 2.99 | <0.001 | 2.05 | 1.57, 2.69 | <0.001 |
| **2nd Quartile PS** | | | | | | |
| Dexamethasone | 0.77 | 0.45, 1.27 | 0.3 | 0.77 | 0.51, 1.13 | 0.2 |
| Age | 1.06 | 1.04, 1.08 | <0.001 | 1.03 | 1.02, 1.04 | <0.001 |
| Q-Score | 1.12 | 1.03, 1.21 | 0.008 | 1.12 | 1.05, 1.20 | <0.001 |
| AST | 1.47 | 1.17, 1.85 | 0.001 | 1.46 | 1.22, 1.74 | <0.001 |
| Creatinine | 1.28 | 1.00, 1.61 | 0.044 | 1.18 | 0.98, 1.42 | 0.080 |
| Platelets | 0.69 | 0.46, 1.04 | 0.074 | 0.80 | 0.59, 1.10 | 0.2 |
| WBC | 1.99 | 1.42, 2.80 | <0.001 | 2.11 | 1.61, 2.76 | <0.001 |
| **3rd Quartile PS** | | | | | | |
| Dexamethasone | 1.24 | 0.78, 1.96 | 0.4 | 1.20 | 0.84, 1.69 | 0.3 |
| Age | 1.06 | 1.04, 1.08 | <0.001 | 1.02 | 1.01, 1.04 | <0.001 |
| Q-Score | 1.13 | 1.02, 1.24 | 0.017 | 1.07 | 0.98, 1.16 | 0.11 |
| AST | 1.36 | 1.10, 1.68 | 0.004 | 1.39 | 1.18, 1.63 | <0.001 |

| | | | | | | |
|---|---|---|---|---|---|---|
| Creatinine | 1.55 | 1.24, 1.94 | <0.001 | 1.36 | 1.13, 1.63 | 0.001 |
| Platelets | 0.77 | 0.53, 1.11 | 0.2 | 0.66 | 0.50, 0.88 | 0.005 |
| WBC | 2.31 | 1.64, 3.28 | <0.001 | 2.55 | 1.95, 3.34 | <0.001 |
| **4th Quartile PS** | | | | | | |
| Dexamethasone | 0.62 | 0.42, 0.90 | 0.014 | 0.61 | 0.44, 0.82 | 0.002 |
| Age | 1.06 | 1.04, 1.07 | <0.001 | 1.02 | 1.01, 1.03 | <0.001 |
| Q-Score | 1.15 | 1.05, 1.25 | 0.002 | 1.07 | 0.99, 1.16 | 0.087 |
| AST | 1.44 | 1.27, 1.64 | <0.001 | 1.36 | 1.22, 1.51 | <0.001 |
| Creatinine | 1.10 | 0.91, 1.33 | 0.3 | 1.20 | 1.02, 1.42 | 0.029 |
| Platelet | 0.65 | 0.50, 0.86 | 0.002 | 0.72 | 0.57, 0.91 | 0.006 |
| WBC | 1.47 | 1.12, 1.94 | 0.006 | 1.59 | 1.26, 2.01 | <0.001 |

[1] OR = Odds Ratio, CI = Confidence Interval. *AST, creatinine, platelet count, and WBC count were log-base-2 transformed.

**Appendix E.** Patients receiving remdesivir: Prediction of in-hospital death/hospice referral and combined in-hospital death/hospice referral and severe outcome by receipt of dexamethasone with logistic regression models, by strata of PS.

| Characteristic* | Death/Hospice | | | Severe or Death/Hospice | | |
|---|---|---|---|---|---|---|
| | OR[1] | 95% CI[1] | p-value | OR[1] | 95% CI[1] | p-value |
| **1st Quartile PS** | | | | | | |
| Dexamethasone | 0.88 | 0.47, 1.65 | 0.7 | 1.02 | 0.62, 1.67 | >0.9 |
| Age | 1.05 | 1.03, 1.08 | <0.001 | 1.02 | 1.00, 1.03 | 0.062 |
| Q-Score | 0.98 | 0.85, 1.11 | 0.7 | 1.02 | 0.91, 1.14 | 0.7 |
| AST | 1.29 | 0.90, 1.87 | 0.2 | 1.26 | 0.95, 1.68 | 0.11 |
| Creatinine | 1.38 | 0.93, 2.03 | 0.10 | 1.12 | 0.81, 1.55 | 0.5 |
| Platelet | 0.50 | 0.28, 0.88 | 0.017 | 0.75 | 0.47, 1.18 | 0.2 |
| WBC | 1.22 | 0.79, 1.85 | 0.4 | 1.54 | 1.09, 2.19 | 0.013 |
| **2nd Quartile PS** | | | | | | |
| Dexamethasone | 0.35 | 0.16, 0.73 | 0.007 | 0.54 | 0.29, 0.98 | 0.048 |
| Age | 1.06 | 1.03, 1.10 | <0.001 | 1.02 | 1.00, 1.05 | 0.042 |
| Q-Score | 1.08 | 0.91, 1.26 | 0.3 | 1.03 | 0.88, 1.18 | 0.7 |
| AST | 1.41 | 0.94, 2.11 | 0.10 | 1.48 | 1.05, 2.09 | 0.025 |
| Creatinine | 1.04 | 0.58, 1.77 | >0.9 | 1.18 | 0.73, 1.85 | 0.5 |
| Platelet | 0.73 | 0.36, 1.48 | 0.4 | 0.50 | 0.27, 0.92 | 0.028 |
| WBC | 1.13 | 0.62, 2.07 | 0.7 | 1.64 | 0.98, 2.79 | 0.064 |
| **3rd Quartile PS** | | | | | | |
| Dexamethasone | 0.76 | 0.37, 1.55 | 0.5 | 0.63 | 0.34, 1.16 | 0.14 |
| Age | 1.08 | 1.05, 1.12 | <0.001 | 1.05 | 1.03, 1.08 | <0.001 |
| Q-Score | 1.12 | 0.96, 1.29 | 0.11 | 1.17 | 1.02, 1.33 | 0.021 |
| AST | 2.04 | 1.32, 3.20 | 0.002 | 2.24 | 1.55, 3.28 | <0.001 |
| Creatinine | 0.55 | 0.28, 1.02 | 0.073 | 0.74 | 0.44, 1.20 | 0.2 |

| | | | | | | |
|---|---|---|---|---|---|---|
| Platelet | 0.72 | 0.37, 1.42 | 0.3 | 0.57 | 0.32, 1.00 | 0.051 |
| WBC | 2.03 | 1.10, 3.79 | 0.024 | 2.48 | 1.45, 4.34 | 0.001 |
| **4th Quartile PS** | | | | | | |
| Dexamethasone | 0.95 | 0.46, 1.96 | 0.9 | 1.21 | 0.66, 2.24 | 0.5 |
| Age | 1.04 | 1.02, 1.08 | 0.004 | 1.02 | 0.99, 1.04 | 0.2 |
| Q-Score | 1.10 | 0.88, 1.34 | 0.4 | 1.12 | 0.92, 1.34 | 0.2 |
| AST | 1.77 | 1.21, 2.59 | 0.003 | 1.89 | 1.37, 2.66 | <0.001 |
| Creatinine | 1.66 | 0.99, 2.76 | 0.051 | 1.21 | 0.75, 1.93 | 0.4 |
| Platelet | 0.47 | 0.23, 0.98 | 0.043 | 0.54 | 0.29, 1.00 | 0.052 |
| WBC | 2.01 | 1.11, 3.67 | 0.022 | 2.65 | 1.61, 4.45 | <0.001 |

[1] OR = Odds Ratio, CI = Confidence Interval. *AST, creatinine, platelet count, and WBC count were log-base-2 transformed.

**Appendix F.** Results of extension of models in table 4a for the non-remdesivir matched group to include quadratic terms for the four log-base-2 transformed laboratory value covariates (creatinine, AST, WBC, and platelet count).

| **Characteristic** | **Death/Hospice** | | | **Severe or Death/Hospice** | | |
|---|---|---|---|---|---|---|
| | **OR**[1] | **95% CI**[1] | **p-value** | **OR**[1] | **95% CI**[1] | **p-value** |
| **Aggregate PS Matched Cohort** | | | | | | |
| Dexamethasone | 0.75 | 0.59, 0.94 | 0.016 | 0.79 | 0.66, 0.95 | 0.013 |
| **1st Quartile PS** | | | | | | |
| Dexamethasone | 0.58 | 0.31, 1.05 | 0.084 | 0.86 | 0.54, 1.34 | 0.5 |
| **2nd Quartile PS** | | | | | | |
| Dexamethasone | 0.75 | 0.44, 1.25 | 0.3 | 0.76 | 0.51, 1.12 | 0.2 |
| **3rd Quartile PS** | | | | | | |
| Dexamethasone | 1.24 | 0.77, 1.97 | 0.4 | 1.20 | 0.84, 1.70 | 0.3 |
| **4th Quartile PS** | | | | | | |
| Dexamethasone | 0.59 | 0.40, 0.86 | 0.008 | 0.57 | 0.41, 0.77 | <0.001 |

[1] OR = Odds Ratio, CI = Confidence Interval

**Appendix G.** Results of extension of models in table 4b for the remdesivir matched group to include quadratic terms for the four log-base-2 transformed laboratory value covariates (creatinine, AST, WBC, and platelet count).

| Characteristic | Death/Hospice | | | Severe or Death/Hospice | | |
|---|---|---|---|---|---|---|
| | OR[1] | 95% CI[1] | p-value | OR[1] | 95% CI[1] | p-value |
| **Aggregate PS Matched Cohort** | | | | | | |
| Dexamethasone | 0.69 | 0.49, 0.97 | 0.034 | 0.79 | 0.60, 1.05 | 0.11 |
| **1st Quartile PS** | | | | | | |
| Dexamethasone | 0.76 | 0.39, 1.46 | 0.4 | 0.88 | 0.52, 1.47 | 0.6 |
| **2nd Quartile PS** | | | | | | |
| Dexamethasone | 0.35 | 0.15, 0.73 | 0.007 | 0.52 | 0.27, 0.95 | 0.036 |
| **3rd Quartile PS** | | | | | | |
| Dexamethasone | 0.73 | 0.35, 1.50 | 0.4 | 0.64 | 0.34, 1.18 | 0.2 |
| **4th Quartile PS** | | | | | | |
| Dexamethasone | 0.98 | 0.47, 2.05 | >0.9 | 1.24 | 0.67, 2.30 | 0.5 |

[1] OR = Odds Ratio, CI = Confidence Interval
